# Nudging Toward Precision Medical Education: Linking Diagnostic Exposures to Tailored Learning

**DOI:** 10.64898/2026.07.31.26358869

**Authors:** Jesse Burk-Rafel, Daniel J. Sartori, Helen Finkelstein, Omar Moussa, Marc M. Triola

**Author notes:** Corresponding Author: Jesse Burk-Rafel, NYU Grossman School of Medicine, 550 1st Avenue, MSB-G110, New York, NY, 10016 United States.

## Abstract

**Problem:** Authentic patient encounters are the raw material of clinical learning, yet the educational resources learners receive are rarely keyed to the diagnoses in front of them, creating temporal and cognitive gaps. Precision medical education (PME) proposes delivering the right resource to the right learner at the right moment, but practical implementation in the clinical learning environment remains limited.

**Approach:** We developed DxMentor, an electronic health record (EHR)-integrated platform that captures each learner’s daily inpatient diagnostic exposures from documented International Classification of Diseases, Tenth Revision (ICD-10) codes. Artificial intelligence (AI) is used to match each diagnosis to an educator-curated formulary of micro-learning resources and board-style questions, and to PubMed-derived primary and synthesis literature converted into plain-language evidence summaries. A personalized email “nudge” is delivered before morning rounds, copying supervising attendings for residents, with engagement tracked longitudinally. We report implementation outcomes from July 2024-April 2026.

**Outcomes:** DxMentor evaluated 32,846 encounters from 335 medical students and 346 internal medicine residents, delivering 17,340 nudges containing 63,754 didactic resources, 17,038 question sets, and 23,594 summarized articles for approximately $390 in AI token costs. In a benchmarking sample, 91.5% (366/400) of diagnosis–resource pairs were rated relevant by physician-educators. Overall, 78.5% (12,393/15,793) of nudges were opened and 11.3% (1,955/17,340) had at least one click. Engagement was higher among residents than students (open: 80.7% vs. 65.8%; click-through: 12.7% vs. 3.1%; both *P* < .001), with substantial between-learner variability.

**Next Steps:** Email opens and clicks are engagement proxies rather than measures of learning. We are therefore linking nudges to educational outcomes, testing alternative recommendation strategies and timing, and expanding to additional specialties and ambulatory and surgical settings.

**Teaser Text:** DxMentor integrates with the electronic health record to capture each learner’s daily diagnostic exposures, then uses AI to match diagnoses with tailored resources and evidence summaries—delivering automated email nudges before rounds to operationalize precision medical education at scale.

## Problem

Authentic patient encounters are the raw material of clinical learning, yet the resources students and residents receive are rarely keyed to the bedside problems in front of them.^1^ Generic clerkship slide decks, board-review platforms, and ad-hoc literature searches position learning asynchronously with clinical decisions, creating temporal and cognitive gaps that can widen as case complexity grows. In one academic year at our institution, medical students and internal medicine residents wrote inpatient notes on over 18,000 patients with more than 4,000 diagnoses—an instructional demand far beyond what even the most dedicated faculty could tailor in real time. This leaves learner-specific gaps hard to identify, separates exam preparation from patient care, and pushes evidence-based medicine (EBM) outside the workflow.

Precision medical education (PME) argues that the convergence of real-time clinical data, learning analytics, and artificial intelligence can deliver “the right resource for the right learner at the right moment.”^2,3^ Yet frictionless implementation of PME innovations into the busy clinical learning environment, without adding burden to learners or faculty, remains undescribed.

Nudges, low-stakes cues, such as a timely email, that make relevant information easy to access without mandating action, have been embedded in electronic health record (EHR) workflows to shape clinician decision-making and improve guideline adherence without adding alert fatigue.^4,5^ In medical education, smaller trials of push-based, just-in-time learning modules have improved question completion, knowledge retention, and perceived relevance relative to static educational resources.^6–8^ Yet these educational nudges are typically delivered days after documentation or outside the primary workspace, leaving the opportunity to connect learning to the moment of clinical care and each learner’s needs unrealized.

## Approach

We created DxMentor, an automated system that detects each learner’s daily diagnostic exposures and uses artificial intelligence (AI) to match diagnoses to curated micro-learning content, board-style questions, and EBM summaries, delivering a nudge within 24 hours. DxMentor was developed iteratively as a quality improvement effort co-produced with students, residents, and faculty. We report design and pilot implementation outcomes from July 1, 2024 to April 30, 2026.

### AI-Searchable Formulary of Micro Educational Resources

We curated an AI-searchable formulary of micro educational resources from institutional subscriptions and free open access medical education (“FOAMed”) sources (**Figure 1A**). These included expert consensus-based didactic articles (expert-authored articles from AMBOSS), diagnostic schemas (The Clinical Problem Solvers), medical education podcasts (The Curbsiders, Core IM), and exam style multiple-choice questions (students: AMBOSS QBank items aligned with the US Medical Licensing Examination; residents: NEJM+ Knowledge questions aligned with the American Board of Internal Medicine blueprint; over 11,000 question items in all). Where possible, resources load automatically via Really Simple Syndication (RSS) feeds and Application Programming Interface (API) queries. As of April 30, 2026, 3,311 distinct resources had been loaded (**Supplemental Material 1**); faculty review the formulary quarterly to keep it current.

**Figure 1.**
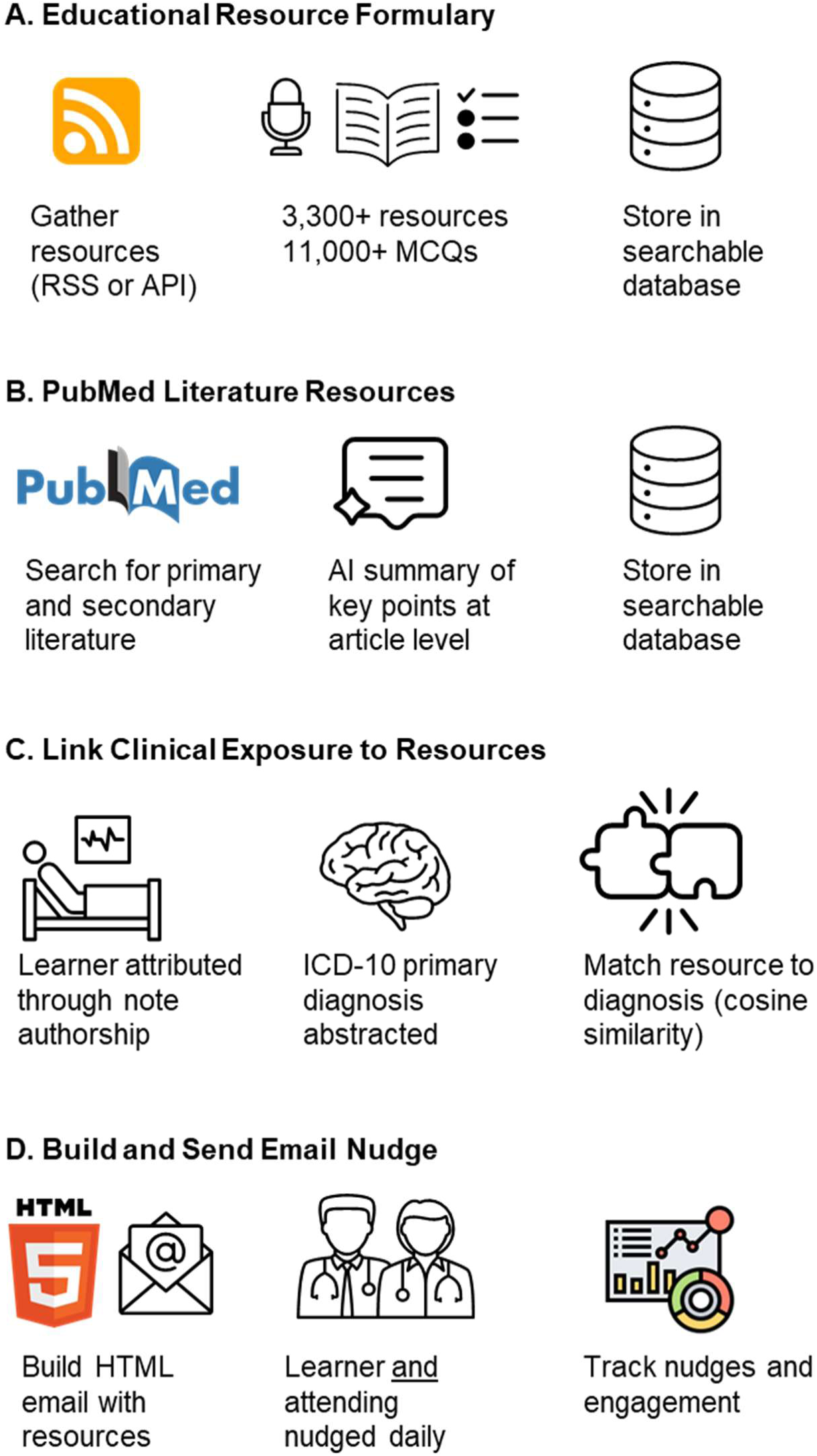
EHR-integrated, AI-powered precision medical education system architecture. (A) An educator-curated formulary of micro-learning resources, including didactic articles, diagnostic schemas, podcasts, and board-style questions, is indexed in a vector database, enabling AI-based semantic search by clinical concept. (B) For each diagnostic encounter, PubMed is queried automatically to retrieve one primary study and one evidence synthesis published within the past five years in approved journals; a generative AI model converts each abstract into a brief plain-language summary. (C) Each learner’s daily diagnostic exposures are captured nightly from the institutional EHR; a rules-based algorithm identifies the most clinically meaningful ICD-10 diagnosis, which is then matched to the most relevant resources in the formulary using AI-calculated semantic similarity. (D) A personalized email nudge assembling the matched micro-learning resources and evidence summaries is delivered to the learner’s inbox before morning rounds, with the supervising attending automatically copied for residents; engagement data (email opens and link clicks) are stored in a longitudinal dashboard accessible to both learners and faculty. Abbreviations: API, Application Programming Interface; HTML, HyperText Markup Language; ICD-10, International Classification of Diseases, Tenth Revision; MCQ, Multiple-Choice Question; RSS, Really Simple Syndication.

Using each resource’s title and descriptive metadata (e.g., podcast episode descriptions), we generated keywords and synonyms for the addressed concepts with generative AI (OpenAI GPT-4.1 v1.0), produced vector representations through an embedding model (nvidia/llama-text-embed-v2), and stored these embeddings in an indexed vector database. Embeddings encode meaning so related terms cluster regardless of wording (“heart attack” maps near “myocardial infarction,” “autoimmune kidney disease” near “lupus nephritis”) enabling DxMentor to retrieve semantically similar resources without keywords (**Supplemental Material 2**).

### Diagnosis-Specific Literature Resources

In addition to micro educational resources, high-quality literature related to each diagnosis is curated (**Figure 1B**). Using the open-source *easy-entrez* library, DxMentor searches PubMed for two articles per diagnostic term: one primary study and one synthesis study. Searches are limited to articles in library-approved journals, English-language, and published within the last five years (journal list and query in **Supplemental Material 3 and 4**). GPT-4.1 is used to convert each abstract into an under 120-word plain-language EBM summary with an institutional library link for paywall-free full-text access.

### Daily Capture of Learner Diagnostic Exposures

DxMentor leverages data from our Epic EHR used at NYU Langone Health Manhattan and Brooklyn (**Figure 1C**). Each morning at 07:00 local time, after Epic’s data warehouse updates, an automated query copies into a dedicated education table every inpatient H&P or progress note written by medical students (core clerkships, electives, and sub-internships) and internal medicine residents. Each row stores the learner’s identifier, note type, the primary and principal International Classification of Diseases, Tenth Revision, Clinical Modification (ICD-10) diagnosis codes recorded in the chart, and the name of any supervising cosigner.

The primary diagnosis is extracted using a rules-based approach that prefers the principal diagnosis selected by the attending chiefly responsible for the patient. Diagnosis codes that denote routine aftercare, screening visits, or procedures are discarded, leaving predominantly ICD-10 codes that reflect diseases.

### Matching Diagnostic Exposures to Micro Educational Resources

For every learner encounter, the AI system calculates a cosine similarity score between the encounter diagnosis and every vector-embedded formulary resource, selecting the most similar (highest scoring) item within each resource category (**Supplemental Material 2**). Diagnoses and resources that have already been suggested to the learner within the past 6 weeks are excluded. This window approximates our clerkship and resident block length, balancing reinforcement against redundancy, consistent with spaced retrieval principles.

### Building and Sending the Nudge

Resources and EBM summaries (with full-text links) are assembled into a mobile-friendly HyperText Markup Language (HTML) email daily at 08:00 local time, arriving just before morning rounds (**Figures 1D, 2**). A single nudge email may span multiple diagnoses.

**Figure 2.**
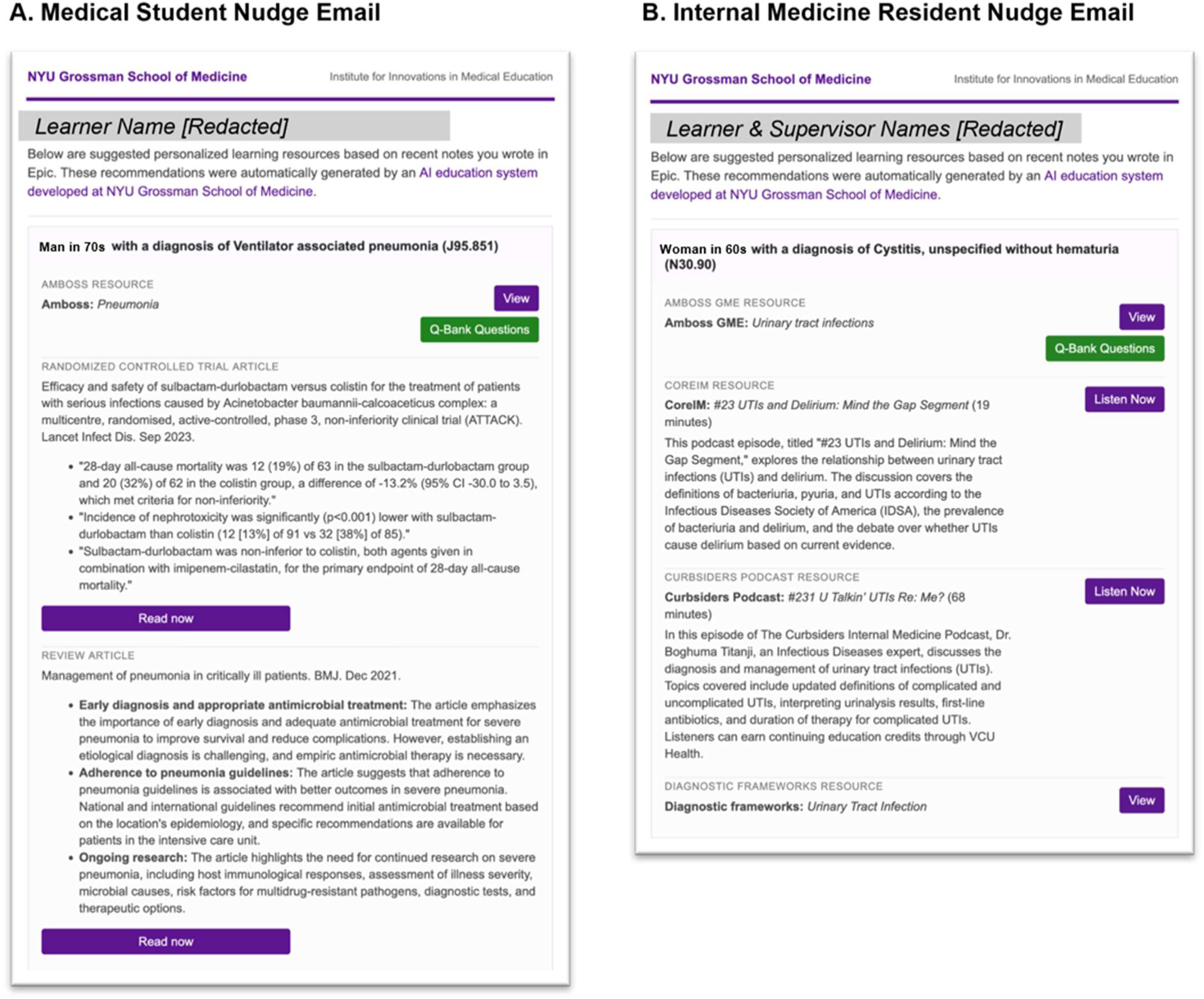
DxMentor EHR-integrated AI-formulated nudge examples. (A) A medical student who wrote a new note for a patient with ventilator-associated pneumonia (ICD-10 code J95.851) receives a nudge with an AMBOSS article and questions on pneumonia, as well as one primary literature article and one review article. (B) An internal medicine resident who wrote a note for a patient with cystitis (ICD-10 code N30.9) receives an AMBOSS GME article and associated NEJM+ knowledge questions on urinary tract infections, two podcasts, and a diagnostic framework.

At faculty request, resident nudges copy the supervising attending (who is attributed to teams quarterly through a human process), reframing the nudge into a tool for collaborative learning, enabling attendings to coach and integrate evidence into rounds. Among medical students, attendings are not copied. For residents, EBM articles are sent weekly as a digest, while students receive them daily.

### Technical Infrastructure and Cost

DxMentor is built in Python using the Django framework and hosted on Red Hat Enterprise Linux 9 web application and MySQL database servers within NYU Langone Health’s secure datacenter environment. The formulary is maintained in a vector database (Pinecone.io), with matching and summarization powered through a cloud-hosted secure generative AI API. No protected health information is included, and all data are encrypted in transit and at rest. Token costs ranged from $0.01 to $0.03 per nudge. DxMentor’s human resource requirements were modest: a physician informaticist (M.M.T.) designed the application and a data engineer (H.F.) maintained the databases.

### Benchmarking and Analysis

We benchmarked the relevance of each AI-generated diagnosis–resource pairing using an ordinal scheme adapted from biomedical information retrieval, in which raters assign degrees of relevance rather than binary labels. A random sample of 400 ICD-10– resource pairs, evenly stratified across resource types, was independently rated by three physician-educators (J.B-R., M.M.T., D.J.S.) on a three-point scale (not relevant; partially relevant; highly relevant); operational definitions and examples are provided in **Supplemental Material 5**. Inter-rater reliability was quantified using quadratically weighted Krippendorf alpha (ɑ*_w_* = 0.60) and agreement (exact agreement across three raters = 64.8%, 259/400; within-one agreement = 97.8%, 391/400). Majority ratings served as the benchmark gold standard; eight pairs had no rater concordance and were resolved through discussion. Matching relevance was then measured against the adjudicated gold standard.

Every nudge, along with whether it was opened (tracking from September 2024) and which links were clicked, is stored in a secure dashboard that learners and faculty can review longitudinally. Group comparisons by learner type were made using chi-square tests, with α set at .05; these analyses were descriptive and did not account for clustering of nudges within learners.

## Outcomes

In a sample of AI pairings of ICD-10 diagnoses and educational resources, 91.5% (366/400, 95% CI 88.4%-93.9%) of pairs were rated either partially or highly relevant (76.5% [306/400] highly relevant; 15.0% [60/400] partially relevant; 8.5% [34/400] not relevant). DxMentor’s diagnosis-to-resource pairing relevance was stronger for some resource modalities, ranging from 100% relevant for question sets and didactic articles to 87.0% for PubMed articles and diagnostic schemas (*P* = .002).

From July 2024 to April 2026, DxMentor emailed 17,340 educational nudges based on 32,846 inpatient encounters attributed to 335 medical students and 346 internal medicine residents (**Table 1**). Medical student nudges averaged 1.2 diagnoses per nudge, while resident nudges averaged 2.1 diagnoses per nudge. Nudges arrived on average 16.7 hours after the attributed note was created. These nudges embedded multiple resources, including 63,754 didactic resource offerings, 17,038 sets of exam-style questions, and 23,594 PubMed articles with AI-generated summaries. The total token cost for generating and summarizing all content was $390, suggesting this AI-supported approach may be scalable at low cost.

**Table 1.** DxMentor nudge volume and engagement among medical students and internal medicine residents at NYU Grossman School of Medicine, July 2024 to April 2026.

|  | **Medical Students**<br>(n=335) | **IM residents**<br>(n=346) | **Overall**<br>(n=681) |
| --- | --- | --- | --- |
| **Patient encounters processed** | 3,154 | 29,692 | 32,846 |
| **Nudge emails delivered** | 2,630 | 14,710 | 17,340 |
| **Nudge emails opened**<br>(proportion, 95% CI)<sup>a</sup> | 1,555<br>(65.8%, 63.8%-67.7%) | 10,838<br>(80.7%, 80.0%-81.4%) | 12,393<br>(78.5%, 77.8%-79.1%) |
| **Click-through count** (proportion, 95% CI) | 82<br>(3.1%, 2.4%-3.8%) | 1,873<br>(12.7%, 12.2%-13.3%) | 1,955<br>(11.3%, 10.8%-11.7%) |
| **Click-through rate among active users,<sup>b</sup> median** (min-max) | 22.2% (3.5%-40.0%) | 29.6% (1.5%-76.5%) | 26.0% (1.5%-76.5%) |

<sup>a</sup> Denominator differs as tracking of email opening became available September 2024; among n=2,365 medical student nudges and n=13,428 resident nudges; adjusted Wald confidence interval.
<sup>b</sup> Calculated for users who received at least 5 nudges and had at least one click, to reflect engagement patterns among the most active user cohorts (n=53 medical students; n=235 residents).

Among nudges sent after email-open tracking was implemented in September 2024, 78.5% (12,393/15,793) were opened; across all nudges, 11.3% (1,955/17,340) had at least one click. This level of engagement is higher than industry educational benchmarks with email open rates around 35% and click rates around 3%.^9^ Click-through rates varied by resource, with diagnostic frameworks having the highest use (**Supplemental Material 6**). Nudge volume differed by learner type: residents received 42.5 nudges per person versus 7.9 for students (*P* < .001), reflecting students’ lower patient volume and shorter inpatient time. Residents had a higher email open rate than students (80.7% vs. 65.8%, *P* < .001) and higher click-through rate (12.7% vs. 3.1%, *P* < .001). These differences should be interpreted cautiously given variation in nudge volume, workflow, and attending copying on resident nudges; usage varied substantially between learners, with a subset of active users driving most clicks.

Among medical students, the top ICD-10 diagnoses associated with nudges reflect note writing across all core clerkships (**Supplemental Material 7**); their top 5 diagnoses were pneumonia (J18.9, 2.5%), unspecified fever (R50.9, 1.8%), acute kidney failure (N17.9, 1.7%), unspecified intestinal obstruction (K56.6, 1.4%), and major depressive disorder (F33.2, 1.3%). Internal medicine residents’ nudges reflected several predominant diagnoses, including pneumonia (J18.9, 5.1%), acute kidney failure (N17.9, 3.7%), heart failure (I50.9, 3.6%), shortness of breath (R06.2, 3.2%), and sepsis (A41.9, 3.0%). The most common literature articles were aligned with these diagnostic areas (**Supplemental Material 8**).

## Next Steps

Nudges were initially envisioned to drive self-regulated learning. To enhance engagement, we are exploring how to incorporate them into team rounds, formative feedback, and coaching. We continue to expand into other settings and are examining deployment in outpatient and surgical environments, which offer unique affordances such as delivering nudges in advance of encounters based on scheduled procedures or visit diagnoses.

Spaced learning (or spaced repetition) has strong evidence of efficacy,^10^ yet the optimal interval and approach in the clinical workspace is unclear. We are therefore comparing nudges on recently seen diagnoses, more distant or never-seen concepts, and each learner’s previously assessed weaknesses. Timing is also highly personal – some learners prefer resources before rounds, others at day’s end – and we are working to customize delivery accordingly.

This innovation has several limitations. First, these implementation outcomes demonstrate feasibility and reach but not educational effectiveness, as email opens and clicks cannot establish resource use or learning; planned qualitative and outcome-linked work will address this. Second, as a single-institution quality improvement effort, our findings may not generalize to other learner populations or EHR settings. Third, ICD-10 diagnoses are a practical but imperfect proxy for diagnostic exposure, as codes may be nonspecific or billing-oriented. Fourth, because benchmarking raters were involved in system design, relevance estimates may be optimistic; future validation will incorporate independent physician-educators. Fifth, copying attendings on resident nudges likely contributed to their higher engagement and creates a tension between formative intent and evaluative perception that warrants further study. Finally, the formulary relies in part on proprietary question banks and a curated, internal medicine-oriented journal list; however, most resources were freely available, and the content-agnostic architecture supports adoption in other contexts.

## Ethical approval

The project was approved as educational quality improvement through an NYU Langone Health IRB certification process.

## Disclaimers

None declared.

## Previous presentations

None declared.

## Data availability

Underlying data falls under HIPAA and FERPA protections and cannot be made available.

## Acknowledgments

The authors wish to thank the students and residents who advised on the creation of this tool. The authors also acknowledge the collaboration of Amboss in sharing a schema for their content catalog. The authors acknowledge librarians at the NYU Health Sciences Library for assistance with formulation of the PubMed search query. The system described in this manuscript is AI-based and incorporates generative AI (GPT-4.1, OpenAI), as detailed in the text. In addition, the authors used AI tools in the preparation of this manuscript. Claude Opus 4.8 (Anthropic), Gemini 2.5 Pro (Google), and GPT-5.4 (OpenAI) were accessed via secure internal instances and used to scaffold an early outline of the manuscript, to revise text for clarity and brevity, and to write code for calculating inter-rater reliability and agreement measures. No primary figures were generated using AI. A human-in-the-loop process was maintained throughout, and the authors reviewed, verified, and edited all AI-assisted content. The authors take full responsibility for the entirety of the manuscript’s contents.

