## Supplemental Materials for "Nudging Toward Precision Medical Education: Linking Diagnostic Exposures to Tailored Learning"

### Supplemental Material

#### Supplemental Material 1: Human curated educational formulary as part of the DxMentor precision medical education platform, as of April 30, 2026

| TYPE | COLLECTION | TARGET LEARNER | DISTINCT ITEMS |
|---|---|---|---|
| Podcasts | The Curbsiders | Resident | 600 |
| Podcasts | Core IM | Resident | 208 |
| Schemas | The Clinical Problem Solvers | Resident | 101 |
| Articles & Multiple-Choice Questions | AMBOSS UME | Medical Student | 1,038[a] |
| Articles & Multiple-Choice Questions | AMBOSS GME | Resident | 1,364 |
| Reviews / Guidelines | PubMed | Medical Student/Resident | 49,818[b] |
| Primary Literature | PubMed | Medical Student/Resident | 45,437[b] |

[a] Number of topics; each topic has multiple MCQs associated totally over 11,000 MCQs
[b] Limited to selected journals, last 5 years, non-retracted adults

#### Supplemental Material 2: Vector database search and logic (generated by Claude Opus 4.8, Anthropic)

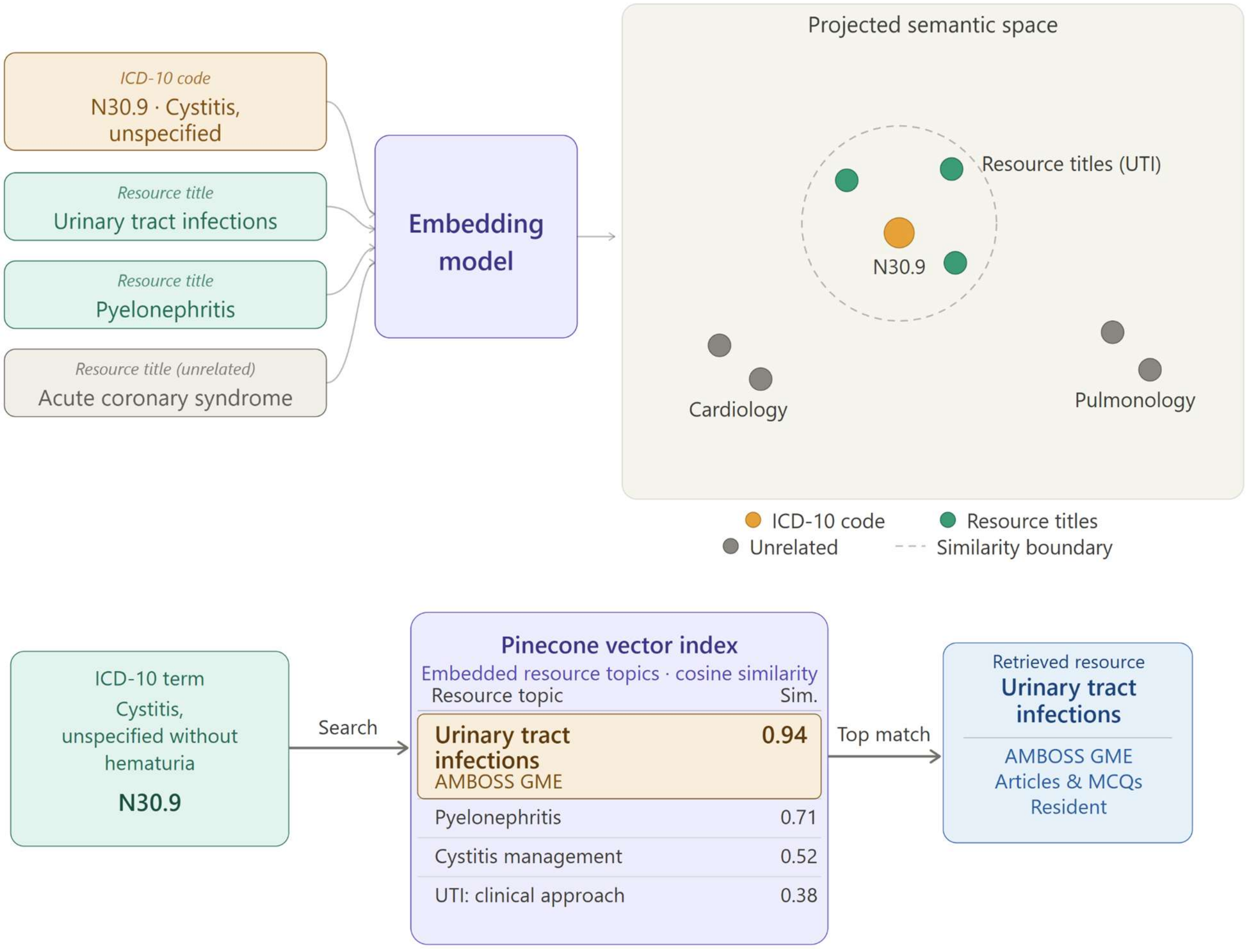


Educational resource index

| Resource | Type | Learner level | |
|---|---|---|---|
| The Curbsiders | Podcast | Resident | |
| Core IM | Podcast | Resident | |
| The Clinical Problem Solvers | Schema | Resident | |
| AMBOSS | Articles & MCQs | Medical student | |
| **AMBOSS GME** | **Articles & MCQs** | **Resident** | matched |

**Supplemental Material 3: Curated PubMed journal list with Medline journal names**

| Number | Journal Name, Medline | Journal Abbreviation, Medline | Target Learner |
|---|---|---|---|
| 1 | Addiction | Addiction | student |
| 2 | Allergy | Allergy | student |
| 3 | American Family Physician | Am Fam Physician | student |
| 4 | American Journal Of Clinical Dermatology | Am J Clin Dermatol | student |
| 5 | American Journal Of Epidemiology | Am J Epidemiol | student |
| 6 | American Journal Of Gastroenterology | Am J Gastroenterol | both |
| 7 | American Journal Of Kidney Diseases | Am J Kidney Dis | resident |
| 8 | American Journal Of Medicine | Am J Med | both |
| 9 | American Journal Of Obstetrics And Gynecology | Am J Obstet Gynecol | student |
| 10 | American Journal Of Ophthalmology | Am J Ophthalmol | student |
| 11 | American Journal Of Psychiatry | Am J Psychiatry | both |
| 12 | American Journal Of Public Health | Am J Public Health | student |
| 13 | American Journal Of Respiratory And Critical Care Medicine | Am J Respir Crit Care Med | both |
| 14 | Annals Of Emergency Medicine | Ann Emerg Med | student |
| 15 | Annals Of Internal Medicine | Ann Intern Med | both |
| 16 | Annals Of The New York Academy Of Sciences | Ann N Y Acad Sci | student |
| 17 | Annals Of Neurology | Ann Neurol | both |
| 18 | Annals Of Oncology | Ann Oncol | both |
| 19 | Antimicrobial Agents And Chemotherapy | Antimicrob Agents Chemother | student |
| 20 | Archives Of Disease In Childhood | Arch Dis Child | student |
| 21 | Archives Of General Psychiatry | Arch Gen Psychiatry | student |
| 22 | Archives Of Internal Medicine | Arch Intern Med | student |
| 23 | Arthritis And Rheumatism | Arthritis Rheum | student |
| 24 | Autoimmunity Reviews | Autoimmun Rev | student |
| 25 | Blood | Blood | both |
| 26 | BMC Cancer | BMC Cancer | student |
| 27 | BMC Medicine | BMC Med | student |
| 28 | BMC Psychiatry | BMC Psychiatry | student |
| 29 | BMC Public Health | BMC Public Health | student |
| 30 | BMJ | BMJ | both |
| 31 | BMJ Open | BMJ Open | student |
| 32 | Brain | Brain | student |
| 33 | Brain Research | Brain Res | student |
| 34 | Cancer Cell | Cancer Cell | student |
| 35 | Cancer Research | Cancer Res | student |
| 36 | Cell | Cell | student |
| 37 | Cell Metabolism | Cell Metab | student |
| 38 | Cell Reports | Cell Rep | student |
| 39 | Chest | Chest | both |
| 40 | Circulation. Arrhythmia And Electrophysiology | Circ Arrhythm Electrophysiol | resident |
| 41 | Circulation Research | Circ Res | both |
| 42 | Circulation | Circulation | both |
| 43 | Clinical Cancer Research | Clin Cancer Res | student |
| 44 | Clinical Diabetes | Clin Diabetes | student |
| 45 | Clinical And Experimental Hypertension | Clin Exp Hypertens | student |
| 46 | Clinical Gastroenterology And Hepatology | Clin Gastroenterol Hepatol | both |
| 47 | Clinical Infectious Diseases | Clin Infect Dis | both |

| | | | |
|---|---|---|---|
| 48 | Clinical Journal Of The American Society Of Nephrology | Clin J Am Soc Nephrol | resident |
| 49 | Clinical Microbiology And Infection | Clin Microbiol Infect | student |
| 50 | Clinical Microbiology Reviews | Clin Microbiol Rev | student |
| 51 | Clinical Psychology Review | Clin Psychol Rev | student |
| 52 | Critical Care Medicine | Crit Care Med | both |
| 53 | Current Opinion In Infectious Diseases | Curr Opin Infect Dis | student |
| 54 | Current Opinion In Virology | Curr Opin Virol | student |
| 55 | Current Psychiatry Reports | Curr Psychiatry Rep | student |
| 56 | Diabetes | Diabetes | student |
| 57 | Diabetes Care | Diabetes Care | student |
| 58 | Diabetes Therapy | Diabetes Ther | student |
| 59 | Drug And Alcohol Dependence | Drug Alcohol Depend | both |
| 60 | Drugs | Drugs | student |
| 61 | Emerging Infectious Diseases | Emerg Infect Dis | student |
| 62 | Endocrine Reviews | Endocr Rev | resident |
| 63 | Endocrinology | Endocrinology | both |
| 64 | Environmental Health Perspectives | Environ Health Perspect | student |
| 65 | Epilepsia | Epilepsia | student |
| 66 | European Heart Journal | Eur Heart J | both |
| 67 | European Journal Of Heart Failure | Eur J Heart Fail | resident |
| 68 | Frontiers In Human Neuroscience | Front Hum Neurosci | student |
| 69 | Frontiers In Immunology | Front Immunol | student |
| 70 | Frontiers In Microbiology | Front Microbiol | student |
| 71 | Frontiers In Neuroscience | Front Neurosci | student |
| 72 | Frontiers In Physiology | Front Physiol | student |
| 73 | Frontiers In Psychology | Front Psychol | student |
| 74 | Gastroenterology | Gastroenterology | both |
| 75 | Gut | Gut | both |
| 76 | Headache | Headache | student |
| 77 | Heart | Heart | student |
| 78 | Hepatology | Hepatology | student |
| 79 | Hypertension | Hypertension | student |
| 80 | Immunity | Immunity | student |
| 81 | Intensive Care Medicine | Intensive Care Med | resident |
| 82 | International Journal Of Hypertension | Int J Hypertens | student |
| 83 | Journal Of Affective Disorders | J Affect Disord | student |
| 84 | Journal Of Allergy And Clinical Immunology | J Allergy Clin Immunol | student |
| 85 | Journal Of The American College Of Cardiology | J Am Coll Cardiol | both |
| 86 | Journal Of The American Geriatrics Society | J Am Geriatr Soc | student |
| 87 | Journal Of The American Medical Association | J Am Med Assoc | both |
| 88 | Journal Of The American Society Of Nephrology | J Am Soc Nephrol | resident |
| 89 | Journal Of Clinical Endocrinology And Metabolism | J Clin Endocrinol Metab | both |
| 90 | Journal Of Clinical Hypertension | J Clin Hypertens | student |
| 91 | Journal Of Clinical Investigation | J Clin Invest | student |
| 92 | Journal Of Clinical Microbiology | J Clin Microbiol | student |
| 93 | Journal Of Clinical Oncology | J Clin Oncol | student |
| 94 | Journal Of Clinical Psychiatry | J Clin Psychiatry | student |
| 95 | Journal Of Diabetes And Its Complications | J Diabetes Complications | student |
| 96 | Journal Of Diabetes Science And Technology | J Diabetes Sci Technol | student |
| 97 | Journal Of General And Family Medicine | J Gen Fam Med | student |
| 98 | Journal Of General Internal Medicine | J Gen Intern Med | both |
| 99 | Journal Of Hematology And Oncology | J Hematol Oncol | resident |

| | | | |
|---|---|---|---|
| 100 | Journal Of Hepatology | J Hepatol | both |
| 101 | Journal Of Hospital Medicine | J Hosp Med | both |
| 102 | Journal Of Hypertension | J Hypertens | student |
| 103 | Journal Of Internal Medicine | J Intern Med | resident |
| 104 | Journal Of The National Cancer Center | J Natl Cancer Cent | resident |
| 105 | Journal Of The National Cancer Institute | J Natl Cancer Inst | student |
| 106 | Journal Of Neurology Neurosurgery And Psychiatry | J Neurol Neurosurg Psychiatry | student |
| 107 | Journal Of Neuroscience | J Neurosci | student |
| 108 | Journal Of Physiology | J Physiol | student |
| 109 | Journal Of Urology | J Urol | student |
| 110 | Journal Of Virology | J Virol | student |
| 111 | JACC. Cardiovascular Imaging | JACC Cardiovasc Imaging | resident |
| 112 | JACC. Cardiovascular Interventions | JACC Cardiovasc Interv | resident |
| 113 | JACC. Heart Failure | JACC Heart Fail | resident |
| 114 | JAMA | JAMA | resident |
| 115 | JAMA Cardiology | JAMA Cardiol | resident |
| 116 | JAMA Dermatology | JAMA Dermatol | resident |
| 117 | JAMA Internal Medicine | JAMA Intern Med | both |
| 118 | JAMA Neurology | JAMA Neurol | resident |
| 119 | JAMA Oncology | JAMA Oncol | resident |
| 120 | JAMA Ophthalmology | JAMA Ophthalmol | resident |
| 121 | JAMA Psychiatry | JAMA Psychiatry | resident |
| 122 | Kidney Medicine | Kidney Med | student |
| 123 | Lancet | Lancet | both |
| 124 | Lancet. Diabetes & Endocrinology | Lancet Diabetes Endocrinol | resident |
| 125 | Lancet. Infectious Diseases | Lancet Infect Dis | both |
| 126 | Lancet. Neurology | Lancet Neurol | both |
| 127 | Lancet. Oncology | Lancet Oncol | both |
| 128 | Lancet. Respiratory Medicine | Lancet Respir Med | resident |
| 129 | Mayo Clinic Proceedings | Mayo Clin Proc | student |
| 130 | Medicine | Medicine (Baltimore) | student |
| 131 | Nature | Nature | student |
| 132 | Nature Biotechnology | Nat Biotechnol | student |
| 133 | Nature Immunology | Nat Immunol | student |
| 134 | Nature Medicine | Nat Med | student |
| 135 | Nature Nanotechnology | Nat Nanotechnol | student |
| 136 | Nature Neuroscience | Nat Neurosci | student |
| 137 | Nature Reviews. Cancer | Nat Rev Cancer | both |
| 138 | Nature Reviews. Cardiology | Nat Rev Cardiol | resident |
| 139 | Nature Reviews. Clinical Oncology | Nat Rev Clin Oncol | resident |
| 140 | Nature Reviews. Drug Discovery | Nat Rev Drug Discov | resident |
| 141 | Nature Reviews. Endocrinology | Nat Rev Endocrinol | both |
| 142 | Nature Reviews. Gastroenterology & Hepatology | Nat Rev Gastroenterol Hepatol | both |
| 143 | Nature Reviews. Immunology | Nat Rev Immunol | resident |
| 144 | Nature Reviews. Microbiology | Nat Rev Microbiol | student |
| 145 | Nature Reviews. Nephrology | Nat Rev Nephrol | resident |
| 146 | Nature Reviews. Neurology | Nat Rev Neurol | resident |
| 147 | Nature Reviews. Neuroscience | Nat Rev Neurosci | both |
| 148 | Nature Reviews. Rheumatology | Nat Rev Rheumatol | resident |
| 149 | Nature Reviews. Urology | Nat Rev Urol | resident |
| 150 | Neuroimage | Neuroimage | both |
| 151 | Neurology | Neurology | student |
| 152 | Neuron | Neuron | student |

| | | | |
|---|---|---|---|
| 153 | Neuropsychiatric Disease And Treatment | Neuropsychiatr Dis Treat | student |
| 154 | Neuroscience And Biobehavioral Reviews | Neurosci Biobehav Rev | student |
| 155 | Neuroscience | Neuroscience | student |
| 156 | New England Journal Of Medicine | N Engl J Med | both |
| 157 | Obstetrics And Gynecology | Obstet Gynecol | student |
| 158 | Ophthalmology | Ophthalmology | student |
| 159 | Palliative Medicine | Palliat Med | resident |
| 160 | Pancreas | Pancreas | student |
| 161 | Pediatrics | Pediatrics | student |
| 162 | PLoS Medicine | PLoS Med | both |
| 163 | PLoS One | PLoS One | student |
| 164 | PLoS Pathogens | PLoS Pathog | student |
| 165 | Primary Care | Prim Care | student |
| 166 | Psychiatry Research | Psychiatry Res | student |
| 167 | Psychological Medicine | Psychol Med | student |
| 168 | Resuscitation | Resuscitation | both |
| 169 | Schizophrenia Bulletin | Schizophr Bull | student |
| 170 | Schizophrenia Research | Schizophr Res | student |
| 171 | Scientific Reports | Sci Rep | student |
| 172 | Science Translational Medicine | Sci Transl Med | student |
| 173 | Science | Science | student |
| 174 | Urology | Urology | student |

**Supplemental Material 4: Example PubMed query on the topic of lobar pneumonia**

("Lobar pneumonia" OR "Pulmonary lobar pneumonia" OR "Pneumonia, Lobar, unspecified organism" OR "Unspecified organism lung infection" OR "Lung lobar pneumonia, no pathogen")

AND (
"Addiction"[Journal] OR "Allergy"[Journal] OR "Am Fam Physician"[Journal] OR "Am J Clin Dermatol"[Journal] OR "Am J Epidemiol"[Journal] OR "Am J Gastroenterol"[Journal] OR "Am J Kidney Dis"[Journal] OR "Am J Med"[Journal] OR "Am J Obstet Gynecol"[Journal] OR "Am J Ophthalmol"[Journal] OR "Am J Psychiatry"[Journal] OR "Am J Public Health"[Journal] OR "Am J Respir Crit Care Med"[Journal] OR "Ann Emerg Med"[Journal] OR "Ann Intern Med"[Journal] OR "Ann N Y Acad Sci"[Journal] OR "Ann Neurol"[Journal] OR "Ann Oncol"[Journal] OR "Antimicrob Agents Chemother"[Journal] OR "Arch Dis Child"[Journal] OR "Arch Gen Psychiatry"[Journal] OR "Arch Intern Med"[Journal] OR "Arthritis Rheum"[Journal] OR "Autoimmun Rev"[Journal] OR "Blood"[Journal] OR "BMC Cancer"[Journal] OR "BMC Med"[Journal] OR "BMC Psychiatry"[Journal] OR "BMC Public Health"[Journal] OR "BMJ"[Journal] OR "BMJ Open"[Journal] OR "Brain"[Journal] OR "Brain Res"[Journal] OR "Cancer Cell"[Journal] OR "Cancer Res"[Journal] OR "Cell"[Journal] OR "Cell Metab"[Journal] OR "Cell Rep"[Journal] OR "Chest"[Journal] OR "Circ Arrhythm Electrophysiol"[Journal] OR "Circ Res"[Journal] OR "Circulation"[Journal] OR "Clin Cancer Res"[Journal] OR "Clin Diabetes"[Journal] OR "Clin Exp Hypertens"[Journal] OR "Clin Gastroenterol Hepatol"[Journal] OR "Clin Infect Dis"[Journal] OR "Clin J Am Soc Nephrol"[Journal] OR "Clin Microbiol Infect"[Journal] OR "Clin Microbiol Rev"[Journal] OR "Clin Psychol Rev"[Journal] OR "Crit Care Med"[Journal] OR "Curr Opin Infect Dis"[Journal] OR "Curr Opin Virol"[Journal] OR "Curr Psychiatry Rep"[Journal] OR "Diabetes"[Journal] OR "Diabetes Care"[Journal] OR "Diabetes Ther"[Journal] OR "Drug Alcohol Depend"[Journal] OR "Drugs"[Journal] OR "Emerg Infect Dis"[Journal] OR "Endocr Rev"[Journal] OR "Endocrinology"[Journal] OR "Environ Health Perspect"[Journal] OR "Epilepsia"[Journal] OR "Eur Heart J"[Journal] OR "Eur J Heart Fail"[Journal] OR "Front Hum Neurosci"[Journal] OR "Front Immunol"[Journal] OR "Front Microbiol"[Journal] OR "Front Neurosci"[Journal] OR "Front Physiol"[Journal] OR "Front Psychol"[Journal] OR "Gastroenterology"[Journal] OR "Gut"[Journal] OR "Headache"[Journal] OR "Heart"[Journal] OR "Hepatology"[Journal] OR "Hypertension"[Journal] OR "Immunity"[Journal] OR "Intensive Care Med"[Journal] OR "Int J Hypertens"[Journal] OR "J Affect Disord"[Journal] OR "J Allergy Clin Immunol"[Journal] OR "J Am Coll Cardiol"[Journal] OR "J Am Geriatr Soc"[Journal] OR "J Am Med Assoc"[Journal] OR "J Am Soc Nephrol"[Journal] OR "J Clin Endocrinol Metab"[Journal] OR "J Clin Hypertens"[Journal] OR "J Clin Invest"[Journal] OR "J Clin Microbiol"[Journal] OR "J Clin Oncol"[Journal] OR "J Clin Psychiatry"[Journal] OR "J Diabetes Complications"[Journal] OR "J Diabetes Sci Technol"[Journal] OR "J Gen Fam Med"[Journal] OR "J Gen Intern Med"[Journal] OR "J Hematol Oncol"[Journal] OR "J Hepatol"[Journal] OR "J Hosp Med"[Journal] OR "J Hypertens"[Journal] OR "J Intern Med"[Journal] OR "J Natl Cancer Cent"[Journal] OR "J Natl Cancer Inst"[Journal] OR "J Neurol Neurosurg Psychiatry"[Journal] OR "J Neurosci"[Journal] OR "J Physiol"[Journal] OR "J Urol"[Journal] OR "J Virol"[Journal] OR "JACC Cardiovasc Imaging"[Journal] OR "JACC Cardiovasc Interv"[Journal] OR "JACC Heart Fail"[Journal] OR "JAMA"[Journal] OR "JAMA Cardiol"[Journal] OR "JAMA Dermatol"[Journal] OR "JAMA Intern Med"[Journal] OR "JAMA Neurol"[Journal] OR "JAMA Oncol"[Journal] OR "JAMA Ophthalmol"[Journal] OR "JAMA Psychiatry"[Journal] OR "Kidney Med"[Journal] OR "Lancet"[Journal] OR "Lancet Diabetes Endocrinol"[Journal] OR "Lancet Infect Dis"[Journal] OR "Lancet Neurol"[Journal] OR "Lancet Oncol"[Journal] OR "Lancet Respir Med"[Journal] OR "Mayo Clin Proc"[Journal] OR "Medicine (Baltimore)"[Journal] OR "N Engl J Med"[Journal] OR "Nat Biotechnol"[Journal] OR "Nat Immunol"[Journal] OR "Nat Med"[Journal] OR "Nat Nanotechnol"[Journal] OR "Nat Neurosci"[Journal] OR "Nat Rev Cancer"[Journal] OR "Nat Rev Cardiol"[Journal] OR "Nat Rev Clin Oncol"[Journal] OR "Nat Rev Drug Discov"[Journal] OR "Nat Rev Endocrinol"[Journal] OR "Nat Rev Gastroenterol Hepatol"[Journal] OR "Nat Rev Immunol"[Journal] OR "Nat Rev Microbiol"[Journal] OR "Nat Rev Nephrol"[Journal] OR "Nat Rev Neurol"[Journal] OR "Nat Rev Neurosci"[Journal] OR "Nat Rev Rheumatol"[Journal] OR "Nat Rev Urol"[Journal] OR

"Nature"[Journal] OR "Neuroimage"[Journal] OR "Neurology"[Journal] OR "Neuron"[Journal] OR "Neuropsychiatr Dis Treat"[Journal] OR "Neurosci Biobehav Rev"[Journal] OR "Neuroscience"[Journal] OR "Obstet Gynecol"[Journal] OR "Ophthalmology"[Journal] OR "Palliat Med"[Journal] OR "Pancreas"[Journal] OR "Pediatrics"[Journal] OR "PLoS Med"[Journal] OR "PLoS One"[Journal] OR "PLoS Pathog"[Journal] OR "Prim Care"[Journal] OR "Psychiatry Res"[Journal] OR "Psychol Med"[Journal] OR "Resuscitation"[Journal] OR "Schizophr Bull"[Journal] OR "Schizophr Res"[Journal] OR "Sci Rep"[Journal] OR "Sci Transl Med"[Journal] OR "Science"[Journal] OR "Urology"[Journal]
)

*AND ("meta-analysis"[Publication Type] OR "review"[Publication Type] OR "systematic review"[Publication Type] OR "guideline"[Publication Type] OR "practice guideline"[Publication Type])*

AND (y_5[Filter]) AND (english[Filter]) AND (humans[Filter])
NOT ("Retracted Publication"[pt] OR "Retraction of Publication"[pt])
NOT (("Adolescent"[Mh] OR "Birth Cohort"[Mh] OR "Child"[Mh] OR "Infant"[Mh] OR "Child, Preschool"[Mh] OR "Infant, Newborn"[Mh]) NOT ("Adult"[Mh]))
NOT (children[ti] OR pediatric[ti])

<u>To search primary literature, replace italicized portion with:</u>
*AND ("clinical study"[Publication Type] OR "clinical trial"[Publication Type] OR "randomized controlled trial"[Publication Type] OR "observational study"[Publication Type] OR "multicenter study"[Publication Type])*

**Supplemental Material 5: Information retrieval rubric for human benchmarking of the relevance of matches between ICD-10 diagnosis codes and educational resource titles**

| Score | Label | Operational definition | Example |
|---|---|---|---|
| 1 | Not Relevant | The educational resource does not meaningfully address the ICD-10 diagnosis, associated condition, management, or core clinical concepts. Any overlap is incidental or overly broad. | ICD-10: E11.9 Type 2 diabetes mellitus without complications<br><br>Resource: “Approach to pediatric asthma exacerbation” |
| 2 | Partially Relevant | The resource includes some content related to the diagnosis or overlapping clinical concepts, but the ICD-10 diagnosis is not a primary focus or coverage is indirect, incomplete, or nonspecific. | ICD-10: I50.9 Heart failure, unspecified<br><br>Resource: “General inpatient fluid management and dyspnea evaluation” |
| 3 | Highly Relevant | The resource directly addresses the ICD-10 diagnosis, including its evaluation, pathophysiology, diagnosis, treatment, complications, or management, and would reasonably support learning related to that condition. | ICD-10: J44.1 COPD with acute exacerbation<br><br>Resource: “Management of acute COPD exacerbations” |

**Supplemental Material 6: Resource distribution count and click-through rates**

| Type | Resource | Number of times distributed | At least one click | Click-through rate |
|---|---|---|---|---|
| Schemas | Diagnostic frameworks | 12,933 | 1,002 | 7.7% |
| Articles & Multiple-Choice Questions | Amboss GME | 13,886 | 761 | 5.5% |
| Literature | PubMed Article | 23,594 | 780 | 3.3% |
| Podcasts | Core IM | 15,133 | 477 | 3.2% |
| Podcasts | Curbsiders Podcast | 18,650 | 496 | 2.7% |
| Articles & Multiple-Choice Questions | Amboss UME | 3,152 | 67 | 2.1% |

**Supplemental Material 7: Top 20 diagnoses among nudges sent to medical students and internal medicine residents from July 2024 to April 2026**

**Top 20 Diagnoses – Medical Students**

| Rank | ICD-10 Diagnosis | ICD-10 Code | Nudge Count | Nudge Proportion[a] |
|---|---|---|---|---|
| 1 | Pneumonia, unspecified organism | J18.9 | 66 | 2.5% |
| 2 | Fever, unspecified | R50.9 | 48 | 1.8% |
| 3 | Acute kidney failure, unspecified | N17.9 | 46 | 1.7% |
| 4 | Unspecified intestinal obstruction, unspecified as to partial versus complete obstruction | K56.609 | 38 | 1.4% |
| 5 | Major depressive disorder, recurrent severe without psychotic features | F33.2 | 35 | 1.3% |
| 6 | Heart failure, unspecified | I50.9 | 34 | 1.3% |
| 7 | Shortness of breath | R06.02 | 34 | 1.3% |
| 8 | Altered mental status, unspecified | R41.82 | 32 | 1.2% |
| 9 | Unspecified abdominal pain | R10.9 | 32 | 1.2% |
| 10 | Sepsis, unspecified organism | A41.9 | 30 | 1.1% |
| 11 | Encounter for full-term uncomplicated delivery | O80.0 | 29 | 1.1% |
| 12 | Weakness | R53.1 | 27 | 1.0% |
| 13 | Acute respiratory failure with hypoxia | J96.01 | 27 | 1.0% |
| 14 | Urinary tract infection, site not specified | N39.0 | 25 | 1.0% |
| 15 | Cerebral infarction, unspecified | I63.9 | 22 | 0.8% |
| 16 | Unspecified convulsions | R56.9 | 21 | 0.8% |
| 17 | Syncope and collapse | R55.0 | 21 | 0.8% |
| 18 | Neutropenia, unspecified | D70.9 | 21 | 0.8% |
| 19 | Single liveborn infant, delivered vaginally | Z38.00 | 21 | 0.8% |
| 20 | Acute cholecystitis | K81.0 | 21 | 0.8% |

[a] Of 2,630 medical student nudges

**Top 20 Diagnoses – Internal Medicine Residents (both daily and weekly nudges)**

| Rank | ICD-10 Diagnosis | ICD-10 Code | Nudge Count | Nudge Proportion[a] |
|---|---|---|---|---|
| 1 | Pneumonia, unspecified organism | J18.9 | 745 | 5.1% |
| 2 | Acute kidney failure, unspecified | N17.9 | 538 | 3.7% |
| 3 | Heart failure, unspecified | I50.9 | 529 | 3.6% |

| | | | | |
|---|---|---|---|---|
| 4 | Shortness of breath | R06.02 | 476 | 3.2% |
| 5 | Sepsis, unspecified organism | A41.9 | 447 | 3.0% |
| 6 | Urinary tract infection, site not specified | N39.0 | 444 | 3.0% |
| 7 | Chest pain, unspecified | R07.9 | 413 | 2.8% |
| 8 | Altered mental status, unspecified | R41.82 | 399 | 2.7% |
| 9 | Anemia, unspecified | D64.9 | 395 | 2.7% |
| 10 | Acute respiratory failure with hypoxia | J96.01 | 377 | 2.6% |
| 11 | Lobar pneumonia, unspecified organism | J18.1 | 358 | 2.4% |
| 12 | Syncope and collapse | R55.0 | 358 | 2.4% |
| 13 | Unspecified atrial fibrillation | I48.91 | 353 | 2.4% |
| 14 | Hypo-osmolality and hyponatremia | E87.1 | 349 | 2.4% |
| 15 | Gastrointestinal hemorrhage, unspecified | K92.2 | 348 | 2.4% |
| 16 | Unspecified fall | W19.0 | 343 | 2.3% |
| 17 | Chronic obstructive pulmonary disease with (acute) exacerbation | J44.1 | 313 | 2.1% |
| 18 | Fever, unspecified | R50.9 | 310 | 2.1% |
| 19 | Non-ST elevation (NSTEMI) myocardial infarction | I21.4 | 288 | 2.0% |
| 20 | Weakness | R53.1 | 261 | 1.8% |

[a] Of 14,710 internal medicine resident nudges

**Supplemental Material 8: Top 20 PubMed articles among nudges sent to medical students and internal medicine residents from July 2024 to April 2026**

**Top 20 Articles – Medical Students**

| Rank | Title | Journal | Publication Date | Nudge Count |
|---|---|---|---|---|
| 1 | 2022 AHA/ACC/HFSA Guideline for the Management of Heart Failure: A Report of the American College of Cardiology/American Heart Association Joint Committee on Clinical Practice Guidelines | Circulation | 2022-05-03 | 51 |
| 2 | Fever of Unknown Origin | N Engl J Med | 2022-02-03 | 42 |
| 3 | Depression | Ann Intern Med | 2021-05-01 | 34 |
| 4 | Hydrocortisone in Severe Community-Acquired Pneumonia | N Engl J Med | 2023-05-25 | 28 |
| 5 | Acute kidney injury | Lancet | 2025-01-18 | 27 |
| 6 | Global, regional, and national incidence and mortality burden of non-COVID-19 lower respiratory infections and aetiologies, 1990-2021: a systematic analysis from the Global Burden of Disease Study 2021 | Lancet Infect Dis | 2024-09-01 | 27 |
| 7 | Efficacy and Safety of AXS-05 (Dextromethorphan-Bupropion) in Patients With Major Depressive Disorder: A Phase 3 Randomized Clinical Trial (GEMINI) | J Clin Psychiatry | 2022-05-30 | 26 |
| 8 | The prevention of perineal trauma during vaginal birth | Am J Obstet Gynecol | 2024-03-01 | 25 |
| 9 | Aggressive or Moderate Fluid Resuscitation in Acute Pancreatitis | N Engl J Med | 2022-09-15 | 24 |
| 10 | Gallstone Disease: Common Questions and Answers | Am Fam Physician | 2024-06-01 | 24 |
| 11 | Community-Acquired Pneumonia | N Engl J Med | 2023-08-17 | 23 |
| 12 | 2021 AHA/ACC/ASE/CHEST/SAEM/SCCT/SCMR Guideline for the Evaluation and Diagnosis of Chest Pain: A Report of the American College of Cardiology/American Heart Association Joint Committee on Clinical Practice Guidelines | Circulation | 2021-11-30 | 22 |
| 13 | Early Restrictive or Liberal Fluid Management for Sepsis-Induced Hypotension | N Engl J Med | 2023-02-09 | 21 |
| 14 | Efficacy and safety of sulbactam-durlobactam versus colistin for the treatment of patients with serious infections caused by Acinetobacter baumannii-calcoaceticus complex: a multicentre, randomised, active-controlled, phase 3, non-inferiority clinical trial (ATTACK) | Lancet Infect Dis | 2023-09-01 | 21 |
| 15 | Combining loop with thiazide diuretics for decompensated heart failure: the CLOROTIC trial | Eur Heart J | 2023-02-01 | 21 |
| 16 | 2024 ACC/AHA/AACVPR/APMA/ABC/SCAI/SVM/SVN/SVS/SIR/VESS Guideline for the Management of Lower Extremity Peripheral Artery Disease: A Report of the American College of Cardiology/American Heart Association Joint Committee on Clinical Practice Guidelines | Circulation | 2024-06-11 | 18 |

| | | | | |
|---|---|---|---|---|
| 17 | Interstitial lung diseases | Lancet | 2022-09-03 | 18 |
| 18 | Compression Therapy to Prevent Recurrent Cellulitis of the Leg | N Engl J Med | 2020-08-13 | 17 |
| 19 | 2023 ACC/AHA/ACCP/HRS Guideline for the Diagnosis and Management of Atrial Fibrillation: A Report of the American College of Cardiology/American Heart Association Joint Committee on Clinical Practice Guidelines | Circulation | 2024-01-02 | 17 |
| 20 | A Randomized Trial of Intravenous Amino Acids for Kidney Protection | N Engl J Med | 2024-08-22 | 17 |

##### Top 20 Articles – IM Residents

| Rank | Title | Journal | Publication Date | Nudge Count |
|---|---|---|---|---|
| 1 | 2022 AHA/ACC/HFSA Guideline for the Management of Heart Failure: A Report of the American College of Cardiology/American Heart Association Joint Committee on Clinical Practice Guidelines | Circulation | 2022-05-03 | 222 |
| 2 | Community-Acquired Pneumonia: A Review | JAMA | 2024-10-15 | 149 |
| 3 | Hydrocortisone in Severe Community-Acquired Pneumonia | N Engl J Med | 2023-05-25 | 138 |
| 4 | 2021 AHA/ACC/ASE/CHEST/SAEM/SCCT/SCMR Guideline for the Evaluation and Diagnosis of Chest Pain: A Report of the American College of Cardiology/American Heart Association Joint Committee on Clinical Practice Guidelines | Circulation | 2021-11-30 | 131 |
| 5 | 2023 ACC/AHA/ACCP/HRS Guideline for the Diagnosis and Management of Atrial Fibrillation: A Report of the American College of Cardiology/American Heart Association Joint Committee on Clinical Practice Guidelines | Circulation | 2024-01-02 | 126 |
| 6 | High-Flow Nasal Oxygen vs Noninvasive Ventilation in Patients With Acute Respiratory Failure: The RENOVATE Randomized Clinical Trial | JAMA | 2025-03-11 | 121 |
| 7 | Compression Therapy to Prevent Recurrent Cellulitis of the Leg | N Engl J Med | 2020-08-13 | 105 |
| 8 | Diagnosis and Management of Hyponatremia: A Review | JAMA | 2022-07-19 | 105 |
| 9 | Urinary Tract Infections: Core Curriculum 2024 | Am J Kidney Dis | 2024-01-01 | 104 |
| 10 | Community-Acquired Pneumonia | N Engl J Med | 2023-08-17 | 99 |
| 11 | Three Weeks Versus Six Weeks of Antibiotic Therapy for Diabetic Foot Osteomyelitis: A Prospective, Randomized, Noninferiority Pilot Trial | Clin Infect Dis | 2021-10-05 | 93 |
| 12 | Prevalence of misdiagnosis of cellulitis: A systematic review and meta-analysis | J Hosp Med | 2023-03-01 | 90 |
| 13 | Risk Assessment and Prevention of Falls in Older Community-Dwelling Adults: A Review | JAMA | 2024-04-23 | 89 |
| 14 | Acute kidney injury | Lancet | 2025-01-18 | 88 |
| 15 | Effect of Regional vs General Anesthesia on Incidence of Postoperative Delirium in Older Patients Undergoing Hip Fracture Surgery: The RAGA Randomized Trial | JAMA | 2022-01-04 | 87 |

| | | | | |
|---|---|---|---|---|
| 16 | Delirium in critical illness: clinical manifestations, outcomes, and management | Intensive Care Med | 2021-10-01 | 87 |
| 17 | Misdiagnosis of Uncomplicated Cellulitis: a Systematic Review and Meta-analysis | J Gen Intern Med | 2023-08-01 | 86 |
| 18 | Prolonged vs Intermittent Infusions of β-Lactam Antibiotics in Adults With Sepsis or Septic Shock: A Systematic Review and Meta-Analysis | JAMA | 2024-08-27 | 81 |
| 19 | ACG Clinical Guideline: Upper Gastrointestinal and Ulcer Bleeding | Am J Gastroenterol | 2021-05-01 | 77 |
| 20 | Cefepime-Taniborbactam in Complicated Urinary Tract Infection | N Engl J Med | 2024-02-15 | 75 |
